# Multiple instance learning using pathology foundation models effectively predicts kidney disease diagnosis and clinical classification

**DOI:** 10.1101/2025.06.03.25328845

**Authors:** Yu Kurata, Imari Mimura, Satoshi Kodera, Hiroyuki Abe, Daisuke Yamada, Haruki Kume, Tetsuo Ushiku, Tetsuhiro Tanaka, Norihiko Takeda, Masaomi Nangaku

## Abstract

**Introduction:** Histological analysis of kidney biopsies is crucial in diagnosing kidney diseases and predicting clinical outcomes. Recently developed pathology foundation models, pretrained on large-scale pathology datasets, have demonstrated excellent performance in various downstream applications. This study evaluated the utility of pathology foundation models combined with multiple instance learning (MIL) for kidney pathology analysis.

**Methods:** We used 242 hematoxylin and eosin (H&E)-stained whole slide images (WSIs) from the Kidney Precision Medicine Project (KPMP) and Japan-Pathology Artificial Intelligence Diagnostics Project (JP-AID) databases as the development cohort, comprising 47 healthy controls, 35 acute interstitial nephritis, and 160 diabetic kidney disease (DKD) slides. External validation was performed using 83 WSIs from the University of Tokyo Hospital (UT dataset). Diagnoses were based on adjudicated diagnoses (KPMP) or expert pathologists-derived diagnoses (JP-AID and UT). Pretrained pathology foundation models were utilized as patch encoders and compared with ImageNet-pretrained ResNet50.

**Results:** In internal validation, all foundation models outperformed ResNet50, achieving area under the receiver operating characteristic curve (AUROC) over 0.980. In external validation, the performance of ResNet50 markedly dropped (AUROC = 0.768), whereas all foundation models maintained higher performance (AUROC over 0.800). Visualization with attention heatmaps confirmed that foundation models accurately recognized diagnostically relevant structures. Additionally, foundation models outperformed ResNet50 in predicting severe proteinuria among DKD cases from KPMP dataset.

**Conclusion:** We successfully integrated pathology foundation models with MIL to achieve robust diagnostic performance, even when trained on a relatively small dataset, highlighting their potential for real-world clinical applications. Key words: artificial intelligence, renal pathology, foundation model, multiple instance learning

## Introduction

The diagnosis of kidney diseases requires interpreting diverse pathological features, such as inflammation, atherosclerosis, and metabolic changes, which demands significant effort and expertise. The advent of whole-slide imaging (WSI) and advances in deep learning have spurred intensive research in digital pathology. In particular, image analysis with convolutional neural networks (CNNs) has been successfully applied to kidney pathology, enabling segmentation of functional structures such as glomeruli and tubules, as well as quantitative analysis of kidney pathology and its association with clinical outcomes^1–3^. These approaches are expected to facilitate efficient kidney histopathological analysis and reduce the time and effort required for evaluation. However, these segmentation-based approaches heavily depend on manually annotated datasets for supervised training, posing a significant limitation to scalability. Self-supervised learning (SSL)^4^ has been introduced as a powerful approach for extracting meaningful representations from unlabeled large datasets. In kidney pathology, the use of SSL for effective feature extraction from glomerular images with limited annotations has been reported^5^. Recently, pathology foundation models, trained using SSL with Vision Transformer (ViT)^6^ architectures, have demonstrated remarkable performance in various downstream tasks compared with ImageNet-trained CNN-based models^7–11^. A key difference between CNNs and ViTs is their inductive biases. CNNs extract spatial patterns using small convolutional kernels across multiple layers and have strong inductive biases, which leads to high performance even with small datasets. However, this inductive bias in turn prevents CNNs from fully leveraging large-scale datasets. In contrast, ViTs have minimal inductive biases, which enable them to outperform CNNs when trained on large-scale datasets^12^. Pathology foundation models can be used as patch-level encoders to extract meaningful features from image patches, and these extracted features can be applied to downstream tasks such as classification and segmentation.

One of the major computational challenges in WSI analysis is the massive data size, making direct slide-level analysis infeasible. The standard solution is dividing each slide into smaller image patches to facilitate computational processing. Multiple instance learning (MIL) is an effective approach for slide-level classification, as it aggregates information from individual patches without requiring patch-level annotations. In heterogeneous tissue slides, where diagnostic value varies widely among patches, MIL excels by learning to focus on important patches for classification.

In this study, we utilized pathology foundation models as encoders to extract features from image patches and employed MIL to aggregate patch-level features for classifying expert pathologist-derived disease categorizations and clinical outcome. Regarding clinical outcome, we evaluated severe proteinuria (albuminuria ≥300 mg/gCre or proteinuria ≥1000 mg/gCre). We then benchmarked the performance of these foundation models against ResNet50^13^, a widely used CNN-based model, to determine their utility in kidney pathology analysis.

## Methods

### Dataset

#### Main experiment

In the main experiment, we developed a diagnostic model for classifying healthy control (HC), acute interstitial nephritis (AIN), and diabetic kidney disease (DKD) using H&E-stained WSIs (Figure 1a). Training data were obtained from two publicly available datasets: the Kidney Precision Medicine Project (KPMP, accessed on 20th January 2025)^14^ and the Japan-Pathology Artificial Intelligence Diagnostics Project (JP-AID, accessed on 20th January 2025)^15^. The KPMP dataset comprised 26 HC slides from 21 patients, 26 AIN slides from 12 patients, and 151 DKD slides from 70 patients. From the JP-AID database, we collected H&E-stained WSIs comprising 21 HC slides, nine DKD slides, and nine AIN slides. For external validation, an independent dataset of H&E-stained WSIs was obtained from biopsy slides collected between 2009 and 2025 at the University of Tokyo Hospital (the UT dataset). The UT dataset included 39 HC slides, 17 AIN slides, and 30 DKD slides. Patients with other coexisting kidney diseases were excluded. Diagnoses were based on adjudicated diagnoses (KPMP) and expert pathologist-derived diagnoses (JP-AID and UT). Most HC cases were derived from transplanted kidney biopsies. Detailed dataset description and selection flowcharts are presented in Supplementary Methods and Figures S1-S3. This study protocol adhered to the Declaration of Helsinki and was approved by the University of Tokyo Institutional Review Board (approval number: 2024526NI).

**Figure 1.**
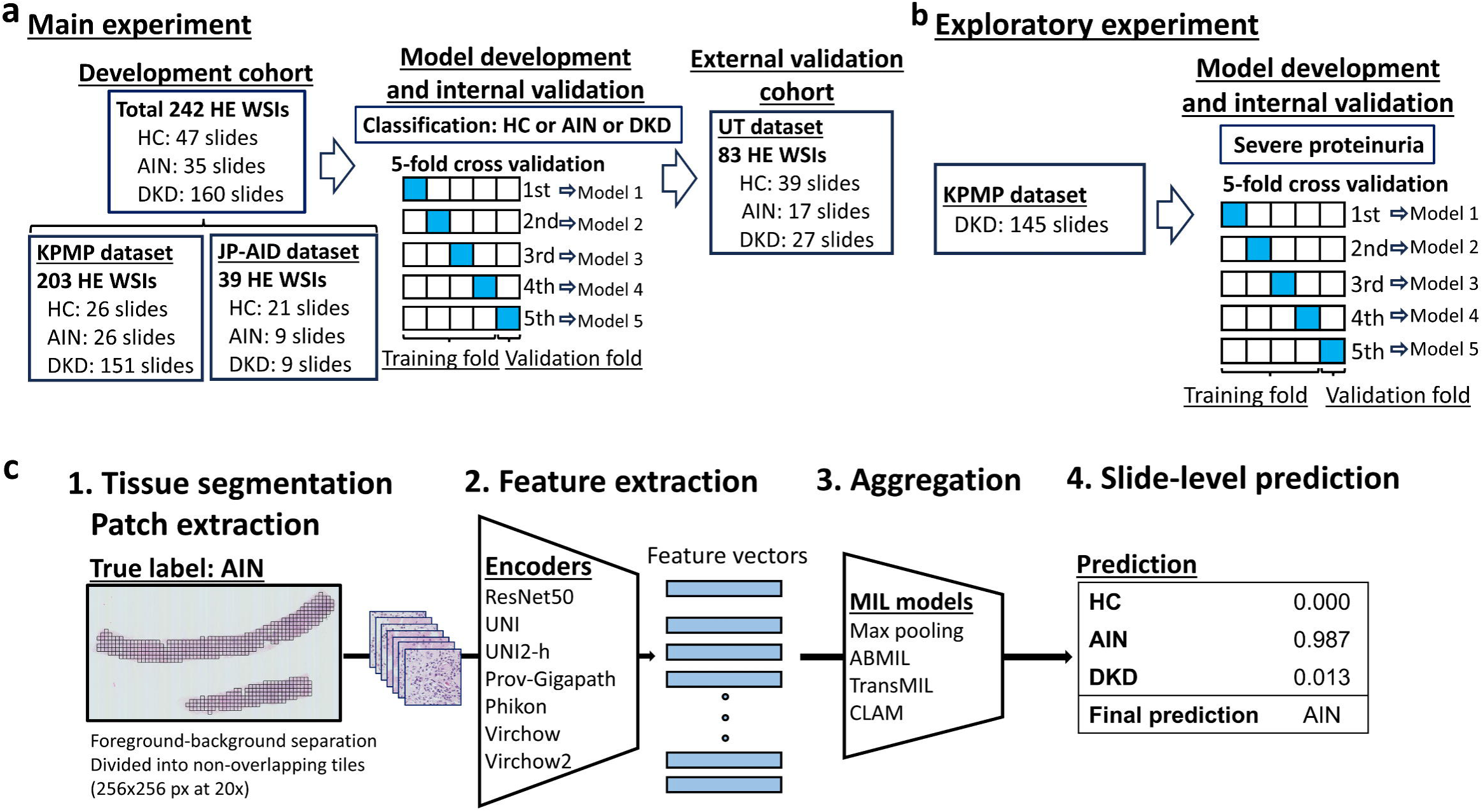
**Overview of study cohorts and workflow of slide-level classification using multiple instance learning** (a) The diagram illustrates the diagnostic model training and evaluation process. The KPMP dataset (total 203 HE WSIs from 103 patients) and the JP-AID dataset (total 39 HE WSIs from 39 patients) across three diagnostic categories were used for model training and internal validation via 5-fold cross validation. The development cohort was randomly split into five subsets while ensuring a balanced distribution of diagnostic categories across the folds. Multiple slides from the same patient were kept in the same subset. In each fold, four subsets were used for training and the remaining subset was used for validation. The 5 trained models were subsequently evaluated on the UT dataset, an external validation cohort comprising 86 H&E-stained WSIs from 86 patients. (b) DKD cases from the KPMP dataset were used for developing the classification model for severe proteinuria in the exploratory experiment. Severe proteinuria was defined as proteinuria ≥1000 mg/gCre or albuminuria ≥300 mg/gCre. For proteinuria classification, six slides from three patients were excluded due to missing data. (c) The workflow for WSI analysis involves four steps: (1) Patch extraction is conducted by dividing WSIs into patches (256×256 pixels) at 20x magnification. Background patches are removed during preprocessing. (2) Feature extraction is performed using pretrained encoders (ResNet50, UNI, UNI2-h, Prov-Gigapath, Phikon, Virchow, and Virchow2). (3) The extracted patch features are aggregated using four MIL methods (max-pooling, ABMIL, TransMIL, and CLAM). (4) The slide-level classifier predicts the diagnostic category. KPMP, kidney precision medicine project; H&E, hematoxylin and eosin; WSIs, whole slide images; JP-AID, Japan-Pathology Artificial Intelligence Diagnostics; HC, healthy control; AIN, acute interstitial nephritis; DKD, diabetic kidney disease; UT, the University of Tokyo Hospital; MIL, multiple instance learning; ABMIL, attention-based MIL; TransMIL, Transformer based MIL; CLAM, clustering-constrained attention multiple instance learning.

#### Exploratory experiment

As an exploratory experiment, we evaluated our pipeline’s ability to predict clinical outcome (severe proteinuria defined as albuminuria ≥300 mg/gCre or proteinuria ≥1000 mg/gCre) using DKD slides from the KPMP dataset (Figure 1b). Six slides from three cases were excluded because of missing values. In this exploratory experiment, external validation using the UT dataset was not performed due to significant class imbalance. Most DKD cases in the UT dataset had severe proteinuria.

#### Image preprocessing

To prepare images for analysis, each WSI was divided into non-overlapping tiles using Slideflow version 3.02^16^ with the Libvips backend at 20x (128 μm, 256 pixels). Background was removed using Otsu’s thresholding^17^, Gaussian blur filtering (sigma = 2, threshold = 0.02), and whitespace filtering (<45% tissue content). Slides containing fewer than 100 patches were excluded to ensure consistent image quality and diagnostic reliability. Analyses were primarily performed without stain normalization, and in the main study we compared performance with and without stain normalization. Thumbnails of representative slides from each dataset are shown in Supplementary Figure S4. Variations in staining protocols and slide scanning methods introduce color heterogeneity, posing challenges in digital pathology analysis^18^. However, recently developed pathology foundation models trained on pathology slides with diverse color variations have demonstrated robust performance even without stain normalization^7, 9, 19, 20^. Given these findings, we opted to primarily analyze the images without stain normalization and assess the robustness of foundation models.

#### Multiple instance learning

For slide-level classification tasks, we adopted the MIL approach. MIL is a weakly supervised learning framework that leverages slide-level labels rather than instance-level labels for learning. In the MIL framework, each WSI is treated as a bag with a slide-level label, whereas the individual patches within the slide that serve as an instance are not labeled. The overall workflow is illustrated in Figure 1c. First, feature vectors were extracted from the extracted patches using pretrained pathology foundation models, including UNI^7^, UNI2-h^7^, Prov-Gigapath^10^, Phikon^11^, Virchow^8^, and Virchow2^9^, which were pretrained on large-scale pathology slides using SSL. As a baseline, we used ResNet50^13^ pretrained on the ImageNet. A summary of the encoders is presented in Table 1. ResNet50 was built from the Torchvision library with ImageNet-pretrained weghts^21^. The pretrained weights of pathology foundation models were downloaded from Huggingface^22^. In this study, we employed four aggregation methods: max pooling, attention-based MIL (ABMIL)^23^, transformer-based MIL (TransMIL)^24^, clustering-constraint attention multiple instance learning (CLAM)^25^. Max pooling assumes that the single most indicative patch in a bag determines the bag label. ABMIL uses an attention mechanism to perform weighted aggregation of patch features, with weights trained by a neural network. TransMIL utilizes a transformer mechanism to learn spatial relationships between patches via self-attention. CLAM incorporates instance-level clustering into its attention mechanism to enhance feature representation learning. Among the two variants of CLAM, we employed the multi-branch variant of CLAM (CLAM-MB), which computes the class-specific attention weights for each class.

**Table 1.**
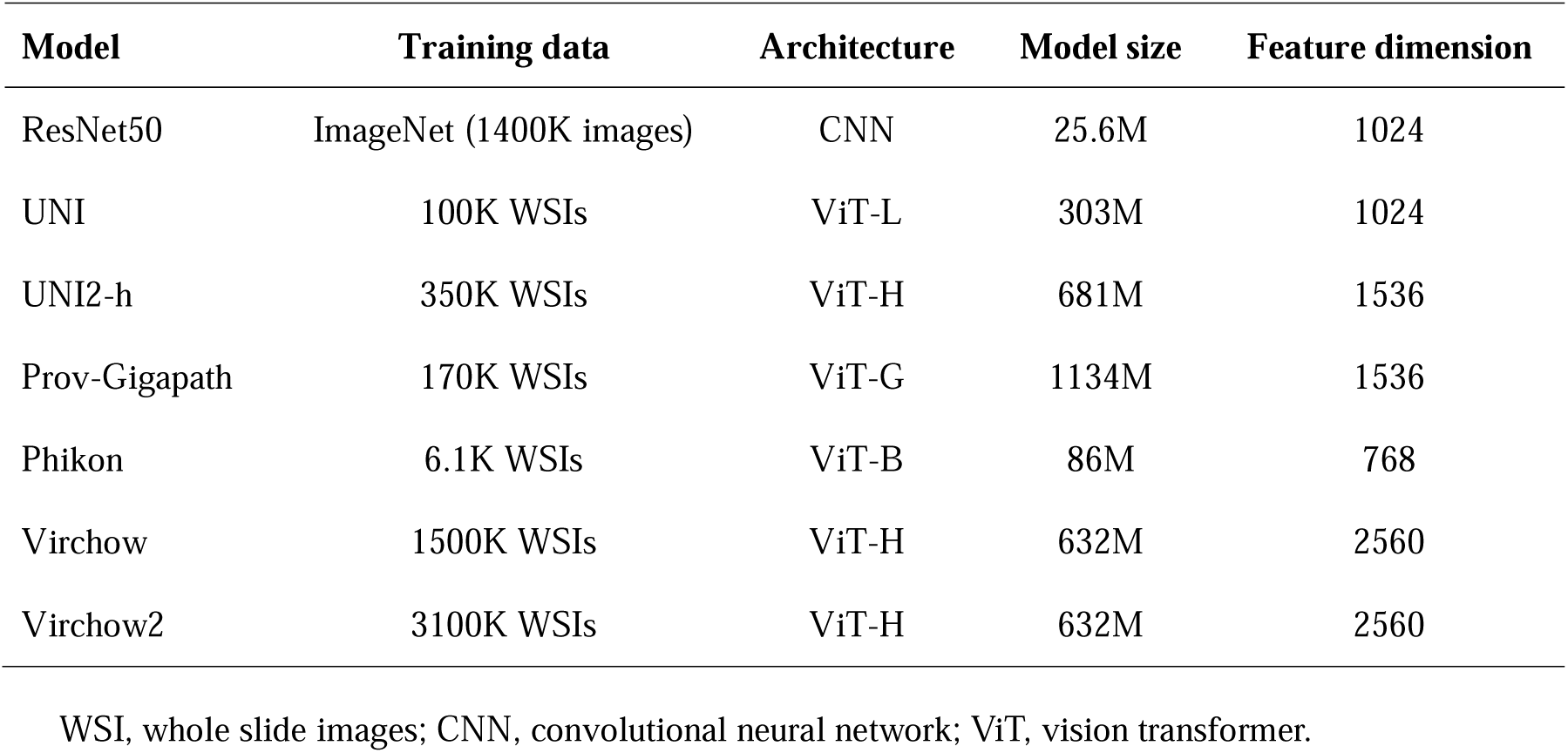
Summary of encoders used in this study. WSI, whole slide images; CNN, convolutional neural network; ViT, vision transformer.

### Model development and validation

#### Main experiment

Each model was trained to predict the diagnostic category among HC, AIN, and DKD. The hyperparameters used for model development are listed in Supplementary Table S1. To develop the model, we performed five-fold cross-validation, in which the dataset was randomly split into five subsets while ensuring a balanced distribution of diagnostic categories across the folds. Multiple slides from the same patient were kept in the same subset. In each fold, four subsets were used for training and the remaining subset was used for validation. This process was repeated five times. The model was trained for 50 epochs per-fold, and the model with the lowest validation loss was selected. The five trained models were subsequently validated using the external UT dataset to assess their generalizability.

#### Exploratory experiment

Five-fold cross validation using DKD cases from the KPMP dataset was performed to predict severe proteinuria. The hyperparameters used in the exploratory experiment were the same as those in the main experiment (Supplementary Table S1). The model was trained for 50 epochs per-fold, and the model with the lowest loss on the internal validation cohort was selected. In this exploratory experiment, aggregation of patch features was performed using CLAM. Visualization Among the four aggregation methods, CLAM-based models were used for visualization. CLAM-MB leverages an attention mechanism to assign a weight to each patch. Based on these attention scores, an attention heatmap is generated to visualize diagnostically relevant regions within the slide. CLAM-MB employs a multi-branch design to learn class-specific attention weights. For each class label, the model generates a distinct attention map that highlights important regions for its classification. We used Slideflow-Studio^16^ to visualize attention maps with the Turbo colormap, where warmer colors (yellow to red) represent higher attention scores while cooler colors (green to blue) indicate lower attention scores. Additionally, we identified the top nine important patches for classification according to their attention scores.

### Statistical analysis

Quantitative or ordinal data are expressed using medians and interquartile ranges or means and 95% confidence intervals (95%CI). Qualitative data are expressed as numbers and percentages. Performance metrics, including accuracy, macro-averaged F1 score, and area under the receiver operating characteristic curve (AUROC) were calculated to assess the predictive performance of the trained models. Accuracy is the proportion of correct classifications out of total classifications. F1 score is the harmonic mean of precision (the proportion of true positive predictions out of all positive predictions) and recall (the proportion of true positive predictions out of all actual positive samples). Macro-averaged F1 score is the arithmetic mean of all per-class F1 scores. The results from the five cross-validation folds were aggregated and reported as the mean and 95%CI from 1000 bootstrap iterations. All metric calculations and statistical analyses were conducted using the scikit-learn package.

## Results

### Main experiment

#### Clinical characteristics of the development cohort

The clinical characteristics of the KPMP and JP-AID cohorts used in the main experiment are presented in Tables 2 and 3. The JP-AID cohort does not provide detailed clinical information other than age, sex, and diagnosis. For eGFR calculations in the KPMP dataset, where eGFR data are provided in ranges (e.g., 40–50 ml/min/1.73 m²), the midpoint eGFR was used (e.g., 45.5 ml/min/1.73 m^2^). For AIN cases in the KPMP cohort, baseline eGFR values and AKI severity were presented. Regarding AKI severity of AIN cases, Kidney Disease: Improving Global Outcomes (KDIGO) AKI stages^26^ were provided, with four patients classified as Stage 2 and eight patients as Stage 3. In the KPMP cohort, DKD cases were characterized by older age and a higher prevalence of hypertension.

**Table 2.**
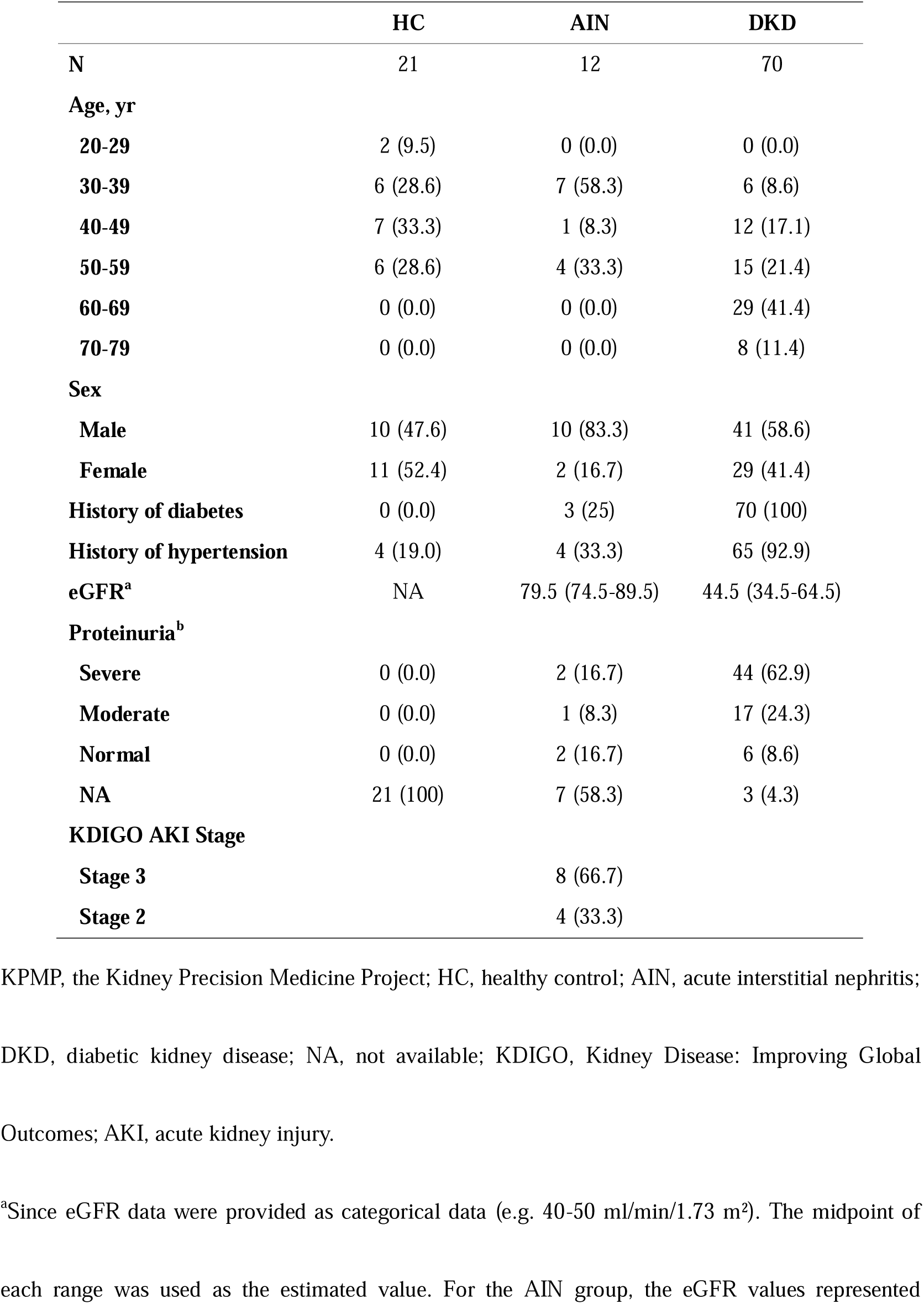
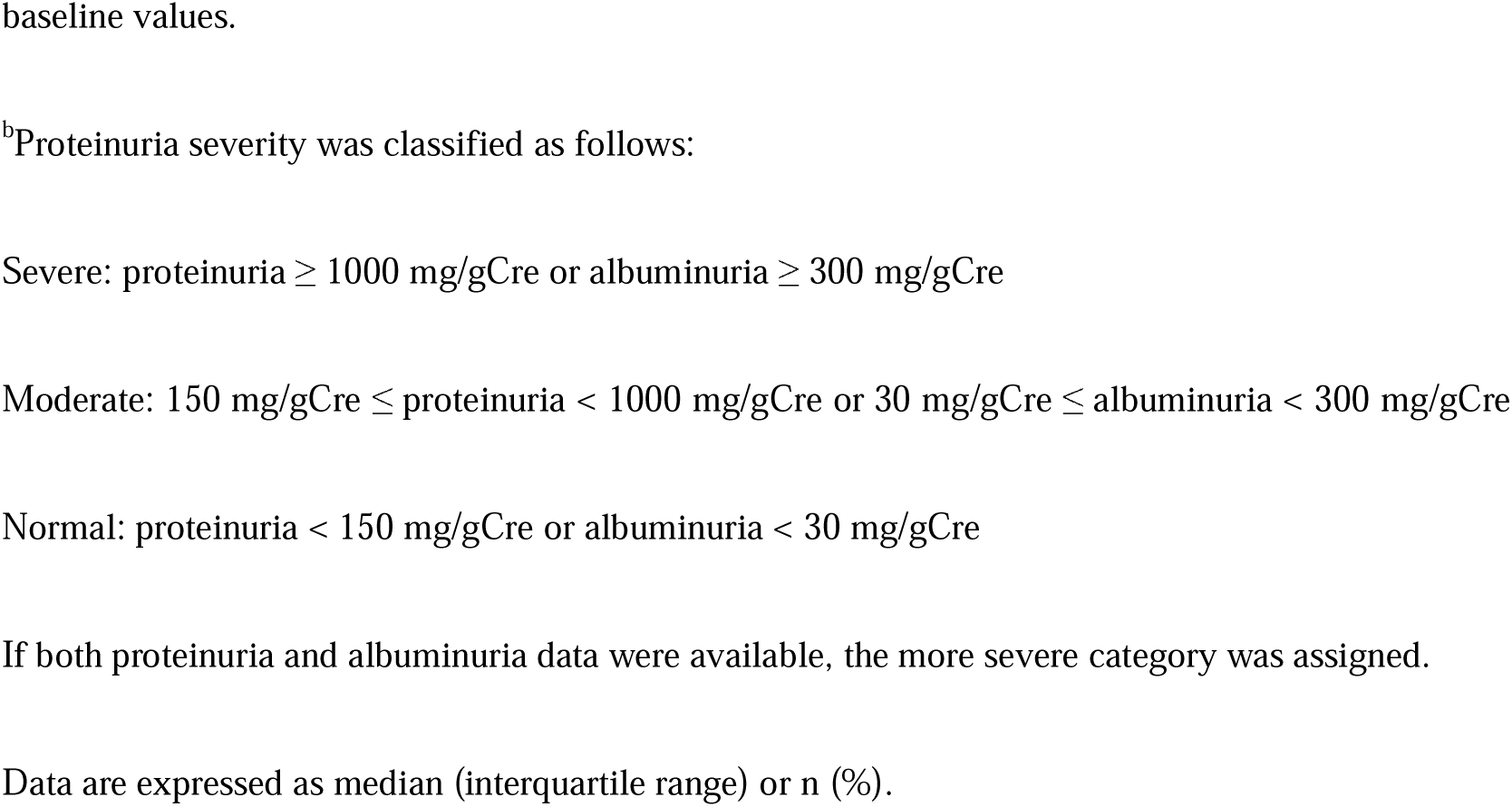
The characteristics of cases in the KPMP cohort.

|  | **HC** | **AIN** | **DKD** |
| --- | --- | --- | --- |
| **N** | 21 | 12 | 70 |
| **Age, yr** |  |  |  |
| **20-29** | 2 (9.5) | 0 (0.0) | 0 (0.0) |
| **30-39** | 6 (28.6) | 7 (58.3) | 6 (8.6) |
| **40-49** | 7 (33.3) | 1 (8.3) | 12 (17.1) |
| **50-59** | 6 (28.6) | 4 (33.3) | 15 (21.4) |
| **60-69** | 0 (0.0) | 0 (0.0) | 29 (41.4) |
| **70-79** | 0 (0.0) | 0 (0.0) | 8 (11.4) |
| **Sex** |  |  |  |
| **Male** | 10 (47.6) | 10 (83.3) | 41 (58.6) |
| **Female** | 11 (52.4) | 2 (16.7) | 29 (41.4) |
| **History of diabetes** | 0 (0.0) | 3 (25) | 70 (100) |
| **History of hypertension** | 4 (19.0) | 4 (33.3) | 65 (92.9) |
| **eGFR<sup>a</sup>** | NA | 79.5 (74.5-89.5) | 44.5 (34.5-64.5) |
| **Proteinuria<sup>b</sup>** |  |  |  |
| **Severe** | 0 (0.0) | 2 (16.7) | 44 (62.9) |
| **Moderate** | 0 (0.0) | 1 (8.3) | 17 (24.3) |
| **Normal** | 0 (0.0) | 2 (16.7) | 6 (8.6) |
| **NA** | 21 (100) | 7 (58.3) | 3 (4.3) |
| **KDIGO AKI Stage** |  |  |  |
| **Stage 3** |  | 8 (66.7) |  |
| **Stage 2** |  | 4 (33.3) |  |

KPMP, the Kidney Precision Medicine Project; HC, healthy control; AIN, acute interstitial nephritis;
DKD, diabetic kidney disease; NA, not available; KDIGO, Kidney Disease: Improving Global
Outcomes; AKI, acute kidney injury.
<sup>a</sup>Since eGFR data were provided as categorical data (e.g. 40-50 ml/min/1.73 m<sup>2</sup>). The midpoint of
each range was used as the estimated value. For the AIN group, the eGFR values represented
baseline values.
<sup>b</sup>Proteinuria severity was classified as follows:
Severe: proteinuria $\geq 1000$ mg/gCre or albuminuria $\geq 300$ mg/gCre
Moderate: $150 \text{ mg/gCre} \leq \text{proteinuria} < 1000 \text{ mg/gCre}$ or $30 \text{ mg/gCre} \leq \text{albuminuria} < 300 \text{ mg/gCre}$

**Table 3.** The characteristics of cases in the JP-AID cohort.

|  | HC<sup>a</sup> | AIN | DKD |
| --- | --- | --- | --- |
| **N** | 21 | 9 | 9 |
| **Age, yr** |  |  |  |
| **10-19** | 1 (4.8) | 2 (22.2) | 0 (0.0) |
| **20-29** | 7 (33.3) | 1 (11.1) | 0 (0.0) |
| **30-39** | 4 (19.0) | 1 (11.1) | 1 (11.1) |
| **40-49** | 3 (14.3) | 0 (0.0) | 0 (0.0) |
| **50-59** | 4 (19.0) | 1 (11.1) | 2 (22.2) |
| **60-69** | 2 (9.5) | 1 (11.1) | 3 (33.3) |
| **70-79** | 0 (0.0) | 1 (11.1) | 4 (44.4) |
| **80-89** | 0 (0.0) | 2 (22.2) | 0 (0.0) |
| **Sex** |  |  |  |
| **Male** | 13 (61.9) | 6 (66.7) | 6 (66.7) |
| **Female** | 8 (38.1) | 3 (33.3) | 3 (33.3) |

JP-AID, Japan-Pathology Artificial Intelligence Diagnostics Project; HC, healthy control; AIN, acute
interstitial nephritis; DKD, diabetic kidney disease.
<sup>a</sup>The demographic information for the transplanted kidneys in the JP-AID HC group may originate
from the recipients rather than the donors. Since the database does not provide detailed clarification,
this remains uncertain and should be interpreted with caution.
Data are expressed as median (interquartile range) or n (%).

#### Model development and internal validation

Based on the development cohort (the KPMP and JP-AID datasets), we developed and evaluated models to predict slide-level diagnosis (HC, AIN, or DKD) using 5-fold cross validation. We performed a comparative evaluation of seven encoders (ResNet50, UNI, UNI2-h, Phikon, Prov-Gigapath, Virchow, Virchow2) with four MIL methods (max pooling, ABMIL, TransMIL, and CLAM). The results are summarized in Figure 2. Among the MIL methods tested, CLAM consistently showed high performance across all seven encoders. In contrast, max pooling underperformed relative to the other MIL methods, although the performance did not decline when Virchow and UNI2-h were used as encoders. Regarding encoders, all pathology foundation models outperformed ResNet50.

**Figure 2.**
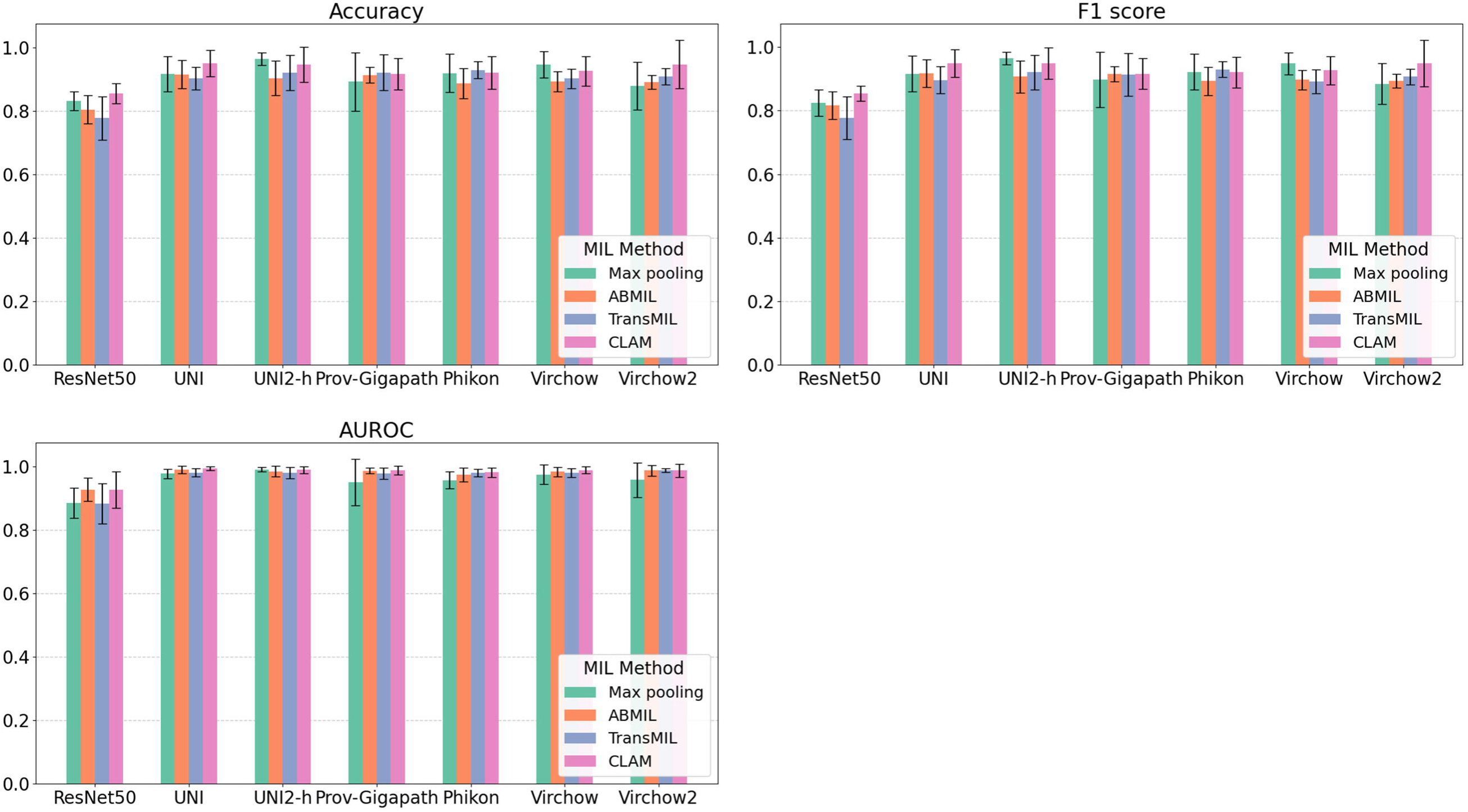
**Model performance for disease classification in the development cohort** Performance metrics (Accuracy, F1 score, and AUROC) of models using seven encoders (ResNet50, UNI, UNI2-h, Prov-Gigapath, Phikon, Virchow, and Virchow2) and four aggregation methods (max pooling, ABMIL, TransMIL, and CLAM) in the internal validation cohort are presented. Data are reported as means and 95% confidence intervals. AUROC, area under the receiver operating characteristic curve; ABMIL, attention-based multiple instance learning; TransMIL, transformer-based multiple instance learning: CLAM, clustering-constraint attention multiple instance learning.

#### External validation

To investigate the robustness and generalizability of the developed models, we assessed their performance on the external validation cohort (the UT dataset). The characteristics of the UT dataset are described in Table 4. Compared with the development cohort, the AIN cases in the UT dataset consisted of older patients, with a greater proportion of females. Additionally, patients with DKD in the UT dataset had lower eGFR levels and a higher prevalence of severe proteinuria. The model performance is summarized in Figure 3. The external validation results followed a similar trend to those observed in the development cohort: CLAM outperformed the other MIL methods, and all pathology foundation models surpassed ResNet50. The performance of max pooling-based models declined substantially in external validation, whereas ABMIL, TransMIL, and CLAM maintained comparable performance. Regarding encoders, ResNet50 exhibited a marked performance decline in the external validation cohort. In contrast, UNI, UNI2-h, Virchow, and Virchow2 maintained high performance. We next assessed the impact of stain variability by applying Macenko stain normalization to all patches^27^. After stain normalization, each model was trained on the development cohort and validated on the external validation cohort using CLAM. As shown in Table 5, stain normalization had a negligible impact on the performance of models based on UNI, UNI2-h, Virchow, and Virchow2, whereas other encoder-based models demonstrated improved performance after stain normalization. These findings indicate the limited generalizability of ImageNet-pretrained ResNet50 and highlight the advantages of pathology foundation models, particularly UNI, UNI2-h, Virchow and Virchow2, for external applicability.

**Figure 3.**
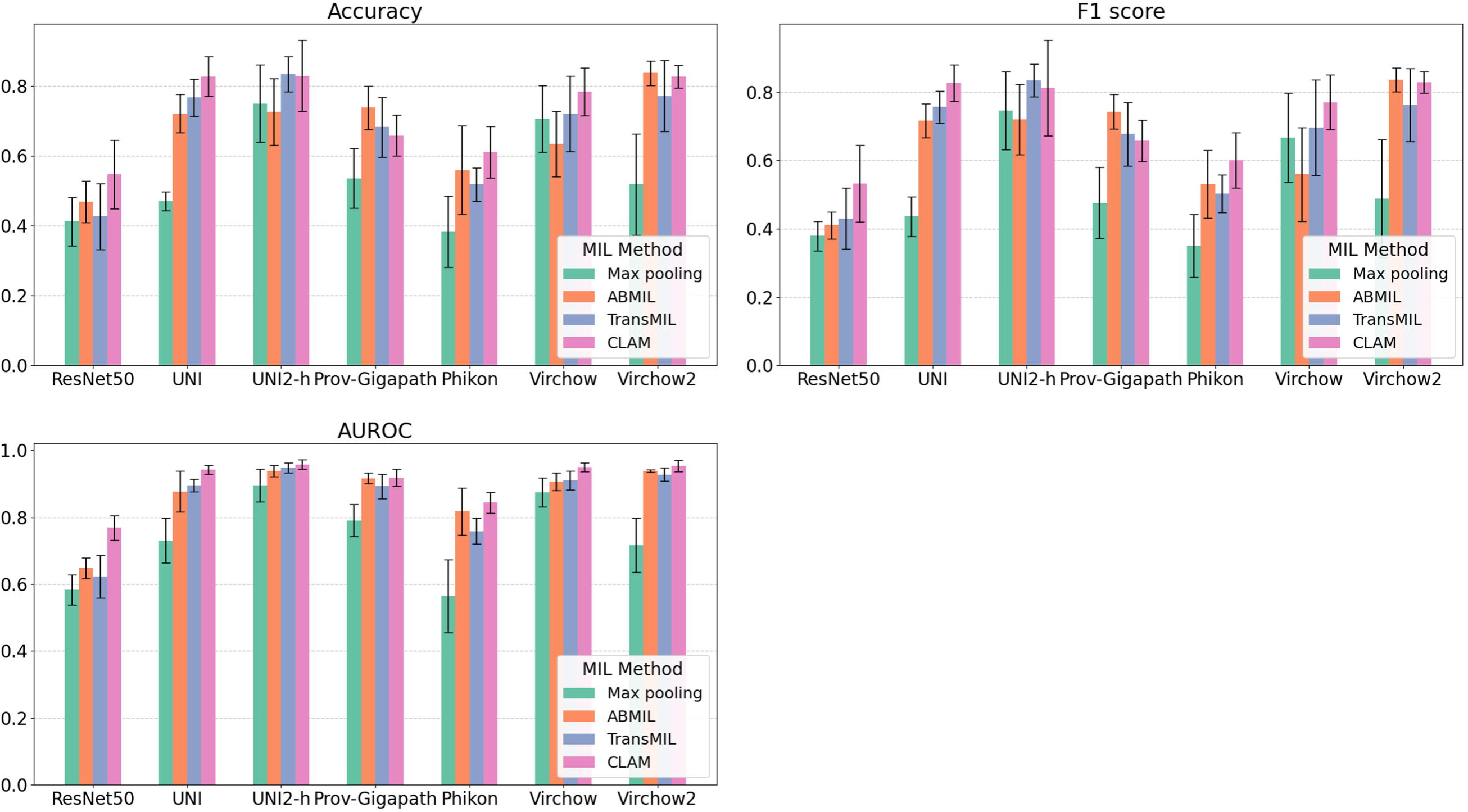
**Model performance for disease classification in the external validation cohort** Performance metrics (Accuracy, F1 score, and AUROC) of models using seven encoders (ResNet50, UNI, UNI2-h, Prov-Gigapath, Phikon, Virchow, and Virchow2) and four aggregation methods (max pooling, ABMIL, TransMIL, and CLAM) in the external validation cohort are presented. Data are reported as means and 95% confidence intervals. AUROC, area under the receiver operating characteristic curve; ABMIL, attention-based multiple instance learning; TransMIL, transformer-based multiple instance learning: CLAM, clustering-constraint attention multiple instance learning.

**Table 4.**
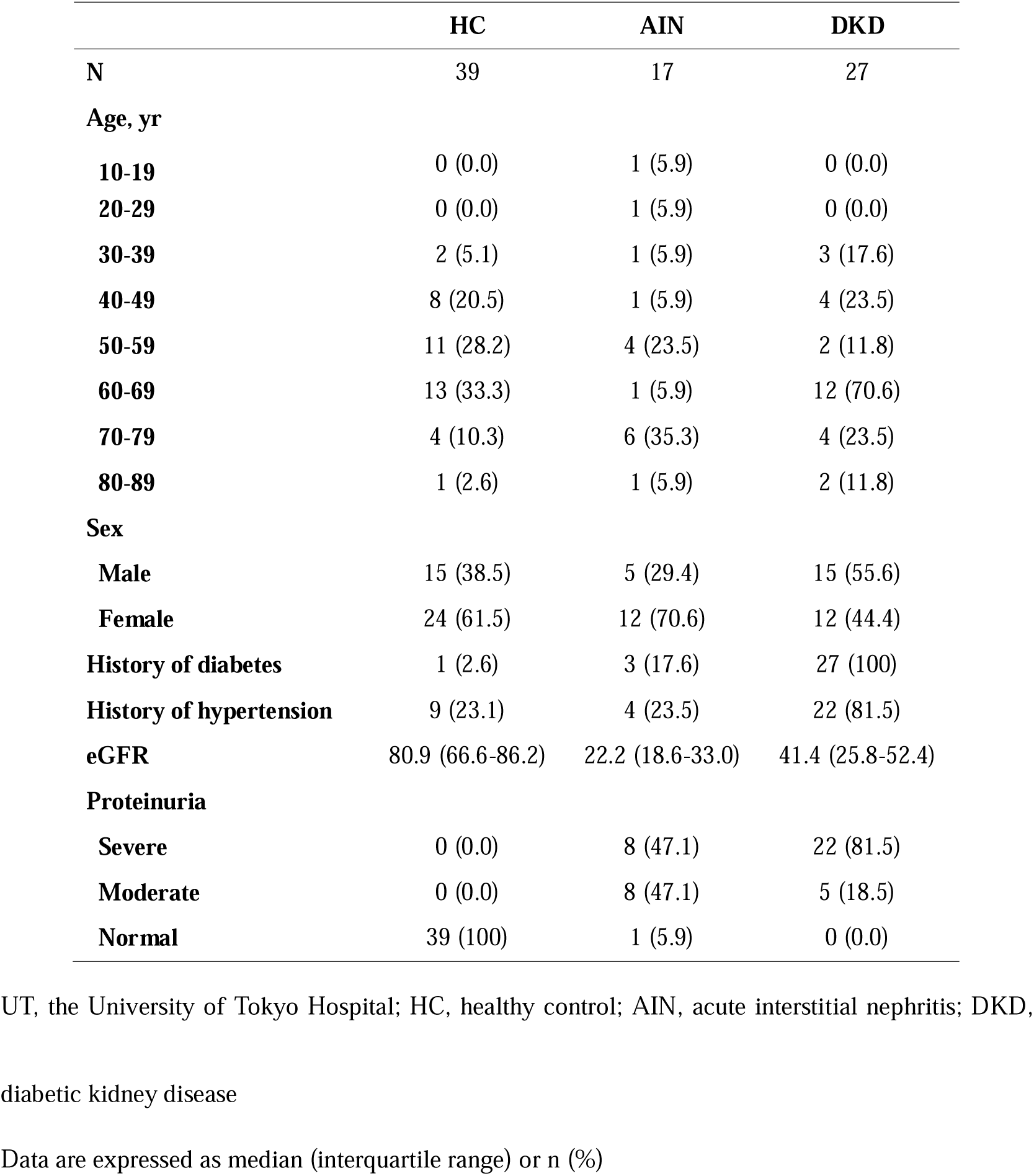
The characteristics of the UT dataset.

**Table 5.** Model performance for disease classification in the validation cohort with or without color normalization.

| **AUROC** | **without stain normalization** | **with stain normalization** |
| --- | --- | --- |
| **ResNet50** | 0.768 (0.740 - 0.798) | 0.815 (0.784 - 0.846) |
| **UNI** | 0.942 (0.933 - 0.953) | 0.945 (0.925 - 0.964) |
| **UNI-2h** | 0.958 (0.945 - 0.967) | 0.966 (0.960 - 0.972) |
| **Prov-Gigapath** | 0.918 (0.896 - 0.935) | 0.944 (0.924 - 0.960) |
| **Phikon** | 0.844 (0.820 - 0.869) | 0.902 (0.883 - 0.926) |
| **Virchow** | 0.949 (0.939 - 0.960) | 0.946 (0.934 - 0.957) |
| **Virchow2** | 0.953 (0.940 - 0.966) | 0.964 (0.959 - 0.969) |

AUROC, area under the receiver operating characteristic curve.
Data are shown in mean (95%CI)

#### Visualization of the developed models

To understand how the developed models identified diagnostically important pathological features, we sampled slides from patients diagnosed with HC, AIN, and DKD in the UT cohort. Using CLAM, which outperformed other MIL methods, we evaluated seven encoders. For each encoder, the best model with the highest AUROC out of five folds in the external validation was used. We created attention heat maps and identified the top nine patches with the highest attention scores per slide (Figure 4). Analysis of these high-attention patches revealed that classification of HC generally relied on recognizing healthy tubules (Figure 4a). Notably, the Phikon-based model did not assign high attention scores to tubular structures, which may explain its inferior performance in the external validation. Interestingly, UNI2-, Prov-Gigapath-, Virchow-, and Virchow2-based models highlighted patches exhibiting coagulative tubular cell necrosis. This is consistent with the fact that HC samples obtained from transplanted kidneys frequently show ischemia-reperfusion injury. Since normal tubules may also appear in varying degrees in AIN and DKD, ischemia-reperfusion injury is more specific to transplanted kidneys. Regarding AIN classification (Figure 4b), all models focused on regions of inflammatory cell infiltration, except for the ResNet50-based model, which incorrectly emphasized largely normal tubules. For diagnosing DKD (Figure 4c), Virchow- and Vichow2-based models specifically focused on glomerular alterations, whereas other encoders highlighted tubulointerstitial and vascular changes. More detailed observations of heat maps from ResNet50- and Virchow2-based models are presented in Figure 5. A sample slide was obtained from a patient diagnosed with AIN in the UT cohort, which is the same slide as shown in Figure 4b. The Virchow2-based model correctly classified this slide as AIN, whereas the ResNet50-based model misclassified it as HC. Attention scores for each diagnostic label (HC, AIN, and DKD) were used to visualize the regions of focus for the models in the disease classification task. In the whole slide view, the Virchow2-based model appropriately captured AIN-related histological features with high attention scores shown in red, covering most of the tissue in the attention map for AIN (Figure 5a). In Region A, where the tissue structure remained relatively intact, the attention map for HC assigned high scores, whereas the attention maps for AIN and DKD assigned low scores. Region B exhibited prominent inflammatory cell infiltration within the interstitium, with high attention scores for AIN and low scores for HC. The attention map for DKD showed elevated scores only in the glomerular regions. These findings indicate that the Virchow2-based model successfully identified key histological patterns associated with each diagnostic category. In contrast, the attention maps generated by the ResNet50-based model failed to capture diagnostically important structures, leading to misclassification of this slide as HC (Figure 5b). While this model correctly assigned high attention scores to Region A for HC classification, it also assigned high scores to Region B, indicating a failure to recognize inflammatory cell infiltration.

**Figure 4.**
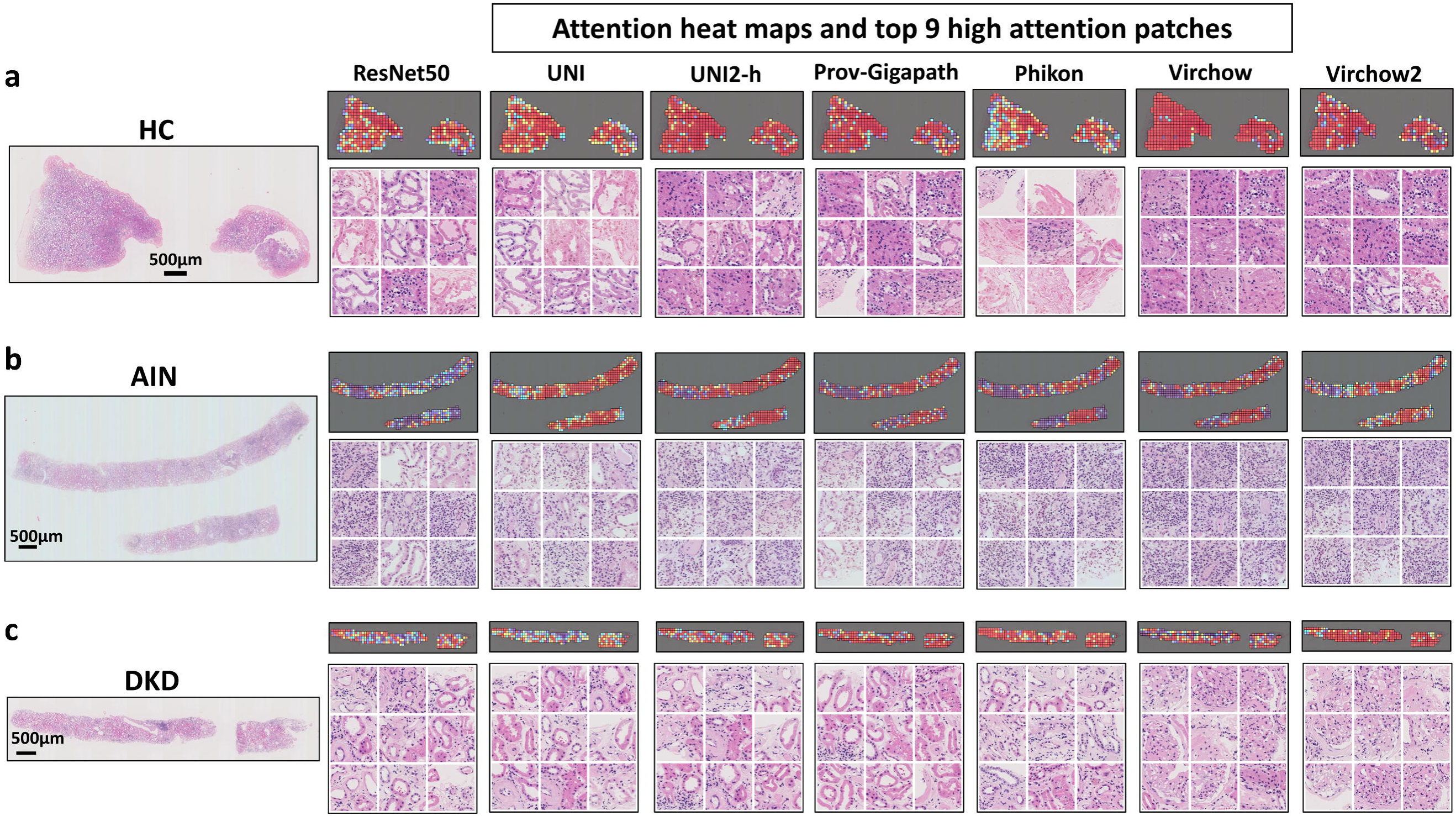
**Attention heatmaps and top nine high-attention patches** HC (a), AIN (b), and DKD (c) slides in the UT dataset were used to visualize how each model built on seven encoders (ResNet50, UNI, UNI2-h, Prov-Gigapath, Phikon, Virchow, and Virchow2) extracted features from image patches. Attention scores for the true label (HC, AIN, or DKD) were used to visualize attention heatmaps with Turbo colormap, where warmer colors (yellow to red) represent higher attention scores, and cooler colors (green to blue) indicate lower attention scores. The top nine patches with the highest attention scores were identified. Scale bar=500 μm. HC, healthy control; AIN, acute interstitial nephritis; DKD, diabetic kidney Disease

**Figure 5.**
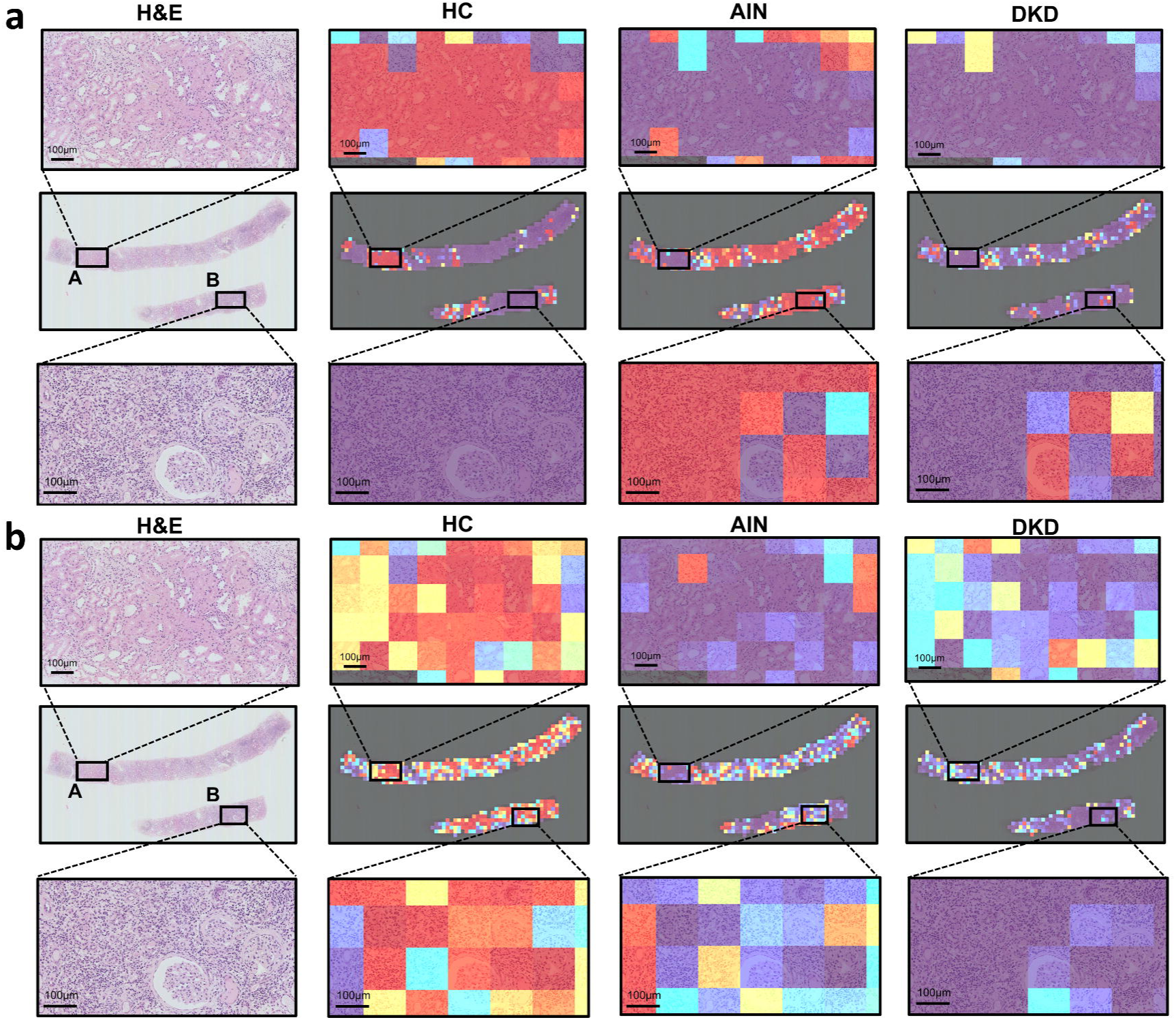
**Attention heatmaps of models using Virchow2 and ResNet50 in the disease classification task** A sample slide was obtained from a patient diagnosed with AIN in the UT cohort. Attention scores for each diagnostic label (HC, AIN, and DKD) were used to visualize the regions of focus for the models in the disease classification task. Heatmaps were generated using the Turbo colormap. Attention heatmaps of the Virchow2-based model (a) and the ResNet50-based model (b) for each label (HC, AIN, and DKD) are shown. The original H&E-stained images were used as references. H&E, hematoxylin-eosin; HC, healthy control; AIN, acute interstitial nephritis; DKD, diabetic kidney disease. Scale bar=100 μm.

#### Exploratory experiment

In the exploratory experiment, we developed predictive models with DKD cases from the KPMP dataset using 5-fold cross-validation, focusing on severe proteinuria (albuminuria ≥300 mg/gCre or proteinuria ≥1000 mg/gCre). In the exploratory experiment, we employed CLAM as the MIL method. The results of the exploratory experiment are summarized in Figure 6. All foundation models showed higher performance than ResNet50, and in particular, UNI2-h demonstrated the highest performance with an AUROC of 0.872 (95%CI 0.794-0.949).

**Figure 6.**
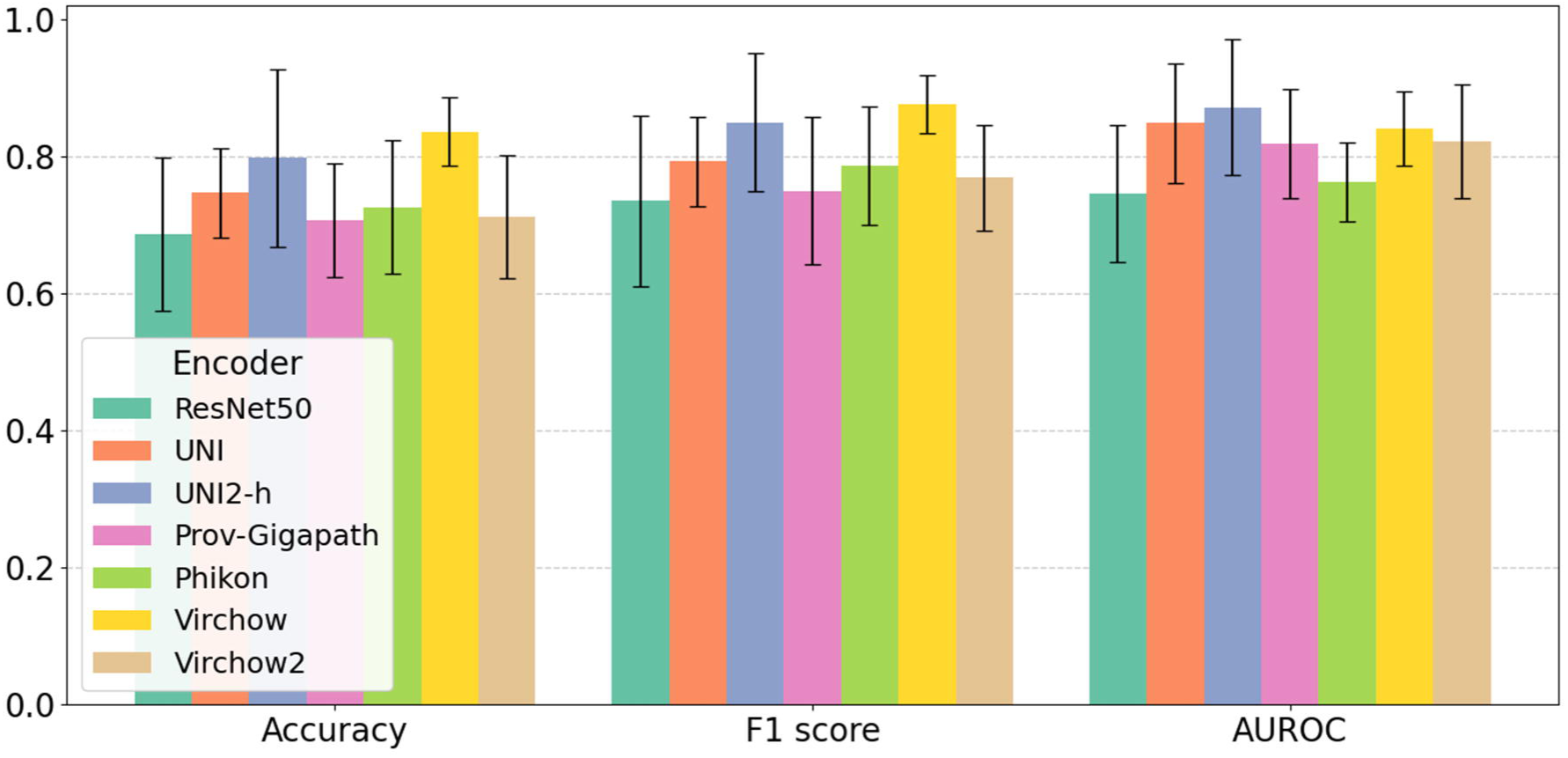
**Model performance for classification of severe proteinuria** Performance metrics (Accuracy, F1 score, and AUROC) of models using seven encoders (ResNet50, UNI, UNI2-h, Prov-Gigapath, Phikon, Virchow, and Virchow2) with CLAM in the KPMP-DKD cohort for the severe proteinuria classification task are presented. Data are reported as means and 95% confidence intervals. AUROC, area under the receiver operating characteristic curve. Data are shown in mean (95%CI)

## Discussion

In this study, we demonstrated that pathology foundation models pretrained on large-scale pathological image datasets are highly effective feature extractors for kidney pathology analysis compared with ResNet50 pretrained on ImageNet. Furthermore, we successfully integrated foundation models with the MIL framework to achieve high diagnostic performance without requiring patch-level annotations. MIL aggregates patch-level features without relying on patch-level labels, making it a practical approach for large-scale histopathological analysis, where detailed manual annotation is infeasible. Among the MIL methods evaluated, CLAM showed the best performance. CLAM aggregates patch-level features through attention pooling and calculates both slide-level classification and instance-level losses to better separate positive and negative instances, enabling the model to efficiently learn discriminative features from important patches. As for disease diagnosis prediction, foundation models’ superiority was consistent when applied to the external validation cohort from a different institution. A major challenge in computational pathology is inter-institutional staining variability, which has been shown to hinder robust feature extraction^28^. Remarkably, our findings indicate that foundation models effectively extracted meaningful features, even without stain normalization, demonstrating robustness against staining variations. This advantage was particularly evident when compared with ResNet50, which exhibited a marked performance drop in external validation. Among the pathology foundation models, UNI, UNI2-h, Virchow, and Virchow2 showed robust performance regardless of whether stain normalization was applied. This finding is consistent with a recent study showing greater robustness of UNI2-h and Virchow2^29^. Attention heat map analysis further revealed that ResNet50 was less effective in extracting pathological features compared with the pathology foundation models. Of note, Virchow and Virchow2 specifically focused on glomeruli in DKD classification, suggesting that these encoders successfully recognized glomerular structures. This finding may be attributed to their substantially larger training datasets compared with other pathology foundation models. Understanding the factors that influence the performance of foundation models in kidney pathology analysis is crucial for optimizing pretraining strategies and developing more effective models. Diagnosis of kidney pathology typically relies on a combination of multiple stains. In this study, we focused on AIN and DKD, both of which exhibit distinct morphological features on H&E-stained slides. This explains why the model for the disease classification task demonstrated a good performance despite being trained solely on H&E-stained slides. For other kidney diseases such as lupus nephritis and membranous glomerulonephritis, integrating multiple stains may be crucial for improving diagnostic accuracy. Indeed, a recent study showed that combining multiple stains enhanced the prediction of clinical remission in lupus nephritis^30^. The foundation models used in this study were trained primarily on H&E slides. Therefore, to improve the diagnostic performance in kidney pathology, developing domain-specific pathology foundation models trained on large-scale, multi-stained slides is an important next step. In the exploratory experiment, we attempted to predict severe proteinuria in patients with DKD using the same pipeline as in the main experiment. Pathology foundation models also demonstrated higher performance than ResNet50. Because the majority of patients with DKD in the UT cohort had severe proteinuria, we could not perform external validation. Therefore, a larger and more diverse external cohort is needed to assess the model’s generalizability. This study has several limitations that warrant consideration. First, post-transplantation cases used as healthy controls exhibited mild ischemia-reperfusion injury, indicating that they may not fully represent normal kidney histology. Second, while we excluded cases with coexisting kidney diseases based on pathology reports, some DKD and AIN slides contained atherosclerotic lesions, likely attributable to aging or hypertension, raising the possibility that the model learns hypertensive changes rather than diabetic changes. In fact, visualization analysis revealed that the models identified patches with interstitial fibrosis as important for DKD classification (Figure 2c). Of note, the models based on Virchow and Virchow2 recognized glomerular alterations as important features for DKD diagnosis, indicating that the pathological features prioritized by the model may differ depending on the encoder. Third, our model was limited to classifying HC, AIN, and DKD. In clinical practice, patients often present with mixed pathological features, complicating categorization into a single diagnosis. Fourth, the relatively small number of slides used in this study may limit the generalizability of our findings. However, even with this limited training set, the pathology foundation models effectively extracted meaningful features and demonstrated good diagnostic performance in the external cohort.

In conclusion, we utilized pathology foundation models as feature extractors and demonstrated superior performance in the slide-level classification of kidney specimens compared with ImageNet-pretrained ResNet50. Their robustness against staining variability and inter-institutional differences highlights their potential for real-world clinical implementation. Since detailed annotations are not required, this approach can be readily extended to diagnosis of other diseases and prediction of clinical outcomes. This approach will facilitate the development of diverse kidney pathology analysis models, which can be particularly valuable in regions with limited access to pathologists and as an educational support tool for trainees.

## Disclosure

All the authors declared no competing interest.

## Data sharing statement

The results here are in part based upon data generated by the Kidney Precision Medicine Project, accessed on January 20, 2025 (https://www.kpmp.org). KPMP is funded by the National Institute of Diabetes and Digestive and Kidney Diseases (Grant numbers: U01DK133081, U01DK133091, U01DK133092, U01DK133093, U01DK133095, U01DK133097, U01DK114866, U01DK114908, U01DK133090, U01DK133113, U01DK133766, U01DK133768, U01DK114907, U01DK114920, U01DK114923, U01DK114933, U24DK114886, UH3DK114926, UH3DK114861, UH3DK114915, UH3DK114937). The KPMP dataset used in this study is publicly available at https://atlas.kpmp.org/. The JP-AID dataset is available at https://jpaid.jp/ upon request, following the prescribed procedures. The UT dataset is not publicly available due to data protection regulations.

## Data Availability

The KPMP dataset used in this study is publicly available at https://atlas.kpmp.org/. The JP-AID dataset is available at https://jpaid.jp/ upon request, following the prescribed procedures. The UT dataset is not publicly available due to data protection regulations.

## Acknowledgement

We sincerely thank Prof. Matthias Kretzler from Division of Computational Medicine and Bioinformatics, University of Michigan, Ann Arbor, Michigan for his helpful advice. We also thank Dr. Isao Matsui from Osaka University, Dr. Ryo Inuzuka, Dr. Masaya Sato, Dr. Hiroaki Ikushima, and Dr. Syohei Hanaoka from the University of Tokyo Hospital for their guidance and insightful discussions that have contributed to the completion of this study. We appreciate Kenichi Hashimoto from Division of Urology of the University of Tokyo in collecting the materials. This research was conducted using the JP-AID database of the Japanese Society of Pathology.

## Author contributions

YK and IM conceived and designed the study. YK, IM, and DY collected clinical data and kidney biopsy specimens. YK, IM, and HA digitized kidney biopsy sections. YK performed data analysis, deep learning, and visualization. Y.K. wrote the manuscript. IM, SK, and TT edited the manuscript. SK, HK, TU, NT, TT, and MN supervised the study.

## Supplementary Material

Supplementary Figure S1 Flowcharts for patients’ inclusion and exclusion in the KPMP dataset

Supplementary Figure S2 Flowcharts for patients’ inclusion and exclusion in the JP-AID dataset

Supplementary Figure S3 Flowcharts for patients’ inclusion and exclusion in the UT dataset

Supplementary Figure S4 Thumbnails of example slides form each dataset

Supplementary Table S1. Hyperparameters for MIL models

Supplementary Methods Dataset description

## References

1. Jayapandian CP, Chen Y, Janowczyk AR, et al. Development and evaluation of deep learning–based segmentation of histologic structures in the kidney cortex with multiple histologic stains. Kidney Int 2021; 99: 86–101.

2. Holscher DL, Bouteldja N, Joodaki M, et al. Next-Generation Morphometry for pathomics-data mining in histopathology. Nat Commun 2023; 14: 470.

3. Pilva P, Bulow R, Boor P. Deep learning applications for kidney histology analysis. Curr Opin Nephrol Hypertens 2024; 33: 291–297.

4. Chen T, Kornblith S, Norouzi M, et al.: A Simple Framework for Contrastive Learning of Visual Representations. In Proceedings of the 37th International Conference on Machine Learning (vol 119), edited by Hal D, III, Aarti S, Proceedings of Machine Learning Research, PMLR, 2020, pp 1597–1607

5. Abe M, Niioka H, Matsumoto A, et al. Self-Supervised Learning for Feature Extraction from Glomerular Images and Disease Classification with Minimal Annotations. J Am Soc Nephrol 2025; 36: 471–486.

6. Dosovitskiy A, Beyer L, Kolesnikov A, et al. An Image is Worth 16×16 Words: Transformers for Image Recognition at Scale. *arXiv* 2020; abs/2010.11929.

7. Chen RJ, Ding T, Lu MY, et al. Towards a general-purpose foundation model for computational pathology. Nat Med 2024; 30: 850–862.

8. Vorontsov E, Bozkurt A, Casson A, et al. Virchow: A Million-Slide Digital Pathology Foundation Model. *arXiv* 2023; abs/2309.07778.

9. Zimmermann E, Vorontsov E, Viret J, et al. Virchow2: Scaling Self-Supervised Mixed Magnification Models in Pathology. *arXiv* 2024; abs/2408.00738.

10. Xu H, Usuyama N, Bagga J, et al. A whole-slide foundation model for digital pathology from real-world data. Nature 2024; 630: 181–188.

11. Filiot A, Ghermi R, Olivier A, et al. Scaling Self-Supervised Learning for Histopathology with Masked Image Modeling. medRxiv 2023.07.21.23292757.

12. Li J, Chen J, Tang Y, et al. Transforming medical imaging with Transformers? A comparative review of key properties, current progresses, and future perspectives. Med Image Anal 2023; 85: 102762.

13. He K, Zhang X, Ren S, et al. Deep Residual Learning for Image Recognition. 2016 IEEE Conference on Computer Vision and Pattern Recognition (CVPR) 2015: 770–778.

14. de Boer IH, Alpers CE, Azeloglu EU, et al. Rationale and design of the Kidney Precision Medicine Project. Kidney Int 2021; 99: 498–510.

15. Sakai Yasuhiro KM, Fukayama Masashi, Yoshizawa Akihiko. Development of artificial intelligence to help pathological diagnosis-Japan pathology AI diagnostics (JP-AID) project. Impact 2019; 6: 40–42.

16. Dolezal JM, Kochanny S, Dyer E, et al. Slideflow: deep learning for digital histopathology with real-time whole-slide visualization. BMC Bioinformatics 2024; 25: 134.

17. Otsu N. A threshold selection method from gray-level histograms. IEEE Trans Syst Man Cybern 1979; 9: 62–66.

18. Schömig-Markiefka B, Pryalukhin A, Hulla W, et al. Quality control stress test for deep learning-based diagnostic model in digital pathology. Mod Pathol 2021; 34: 2098–2108.

19. Vorontsov E, Bozkurt A, Casson A, et al. A foundation model for clinical-grade computational pathology and rare cancers detection. Nat Med 2024; 30: 2924–2935.

20. Wolflein G, Ferber D, Meneghetti AR, et al. Benchmarking Pathology Feature Extractors for Whole Slide Image Classification. arXiv 2024; arXiv:2311.11772v5.

21. TorchVision: PyTorch’s Computer Vision library. In, GitHub repository, GitHub, 2016

22. Wolf T, Debut L, Sanh V, et al. HuggingFace’s Transformers: State-of-the-art Natural Language Processing. arXiv 2019; **abs/**1910**.03771**.

23. Maximilian Ilse JMT, Max Welling. Attention-based Deep Multiple Instance Learning. arXiv 2018; 1802.04712v4.

24. Zhuchen Shao HB, Yang Chen, Yifeng Wang, Jian Zhang, Xiangyang Ji, Yongbing Zhang. TransMIL: Transformer based Correlated Multiple Instance Learning for Whole Slide Image Classification. arXiv 2021; 2106.00908v2.

25. Lu MY, Williamson DFK, Chen TY, et al. Data-efficient and weakly supervised computational pathology on whole-slide images. Nat Biomed Eng 2021; 5: 555–570.

26. KDIGO Clinical Practice Guideline for Acute Kidney Injury. Kidney Int Suppl 2012; 2.

27. Macenko M, Niethammer M, Marron JS, et al. A method for normalizing histology slides for quantitative analysis. 2009 IEEE International Symposium on Biomedical Imaging: From Nano to Macro 2009: 1107–1110.

28. Boschman J, Farahani H, Darbandsari A, et al. The utility of color normalization for AI-based diagnosis of hematoxylin and eosin-stained pathology images. J Pathol 2022; 256: 15–24.

29. Edwin D. de Jong EM, Jonas Teuwen. Current Pathology Foundation Models are unrobust to Medical Center Differences. arXiv 2025; **arXiv:**2501**.18055**.

30. Cheng C, Li B, Li J, et al. Multi-stain deep learning prediction model of treatment response in lupus nephritis based on renal histopathology. Kidney Int 2025; 107: 714–727.

