## Supplementary for "Multiple instance learning using pathology foundation models effectively predicts kidney disease diagnosis and clinical classification"

**Supplementary material**

| | |
|---|---|
| **Supplementary Figure S1** | Flowcharts for patients' inclusion and exclusion in the KPMP dataset |
| **Supplementary Figure S2** | Flowcharts for patients' inclusion and exclusion in the JP-AID dataset |
| **Supplementary Figure S3** | Flowcharts for patients' inclusion and exclusion in the UT dataset |
| **Supplementary Figure S4** | Thumbnails of example slides form each dataset |
| **Supplementary Table S1** | Hyperparameters for MIL models |
| **Supplementary Methods** | Dataset description |

**Supplementary Figure S1**

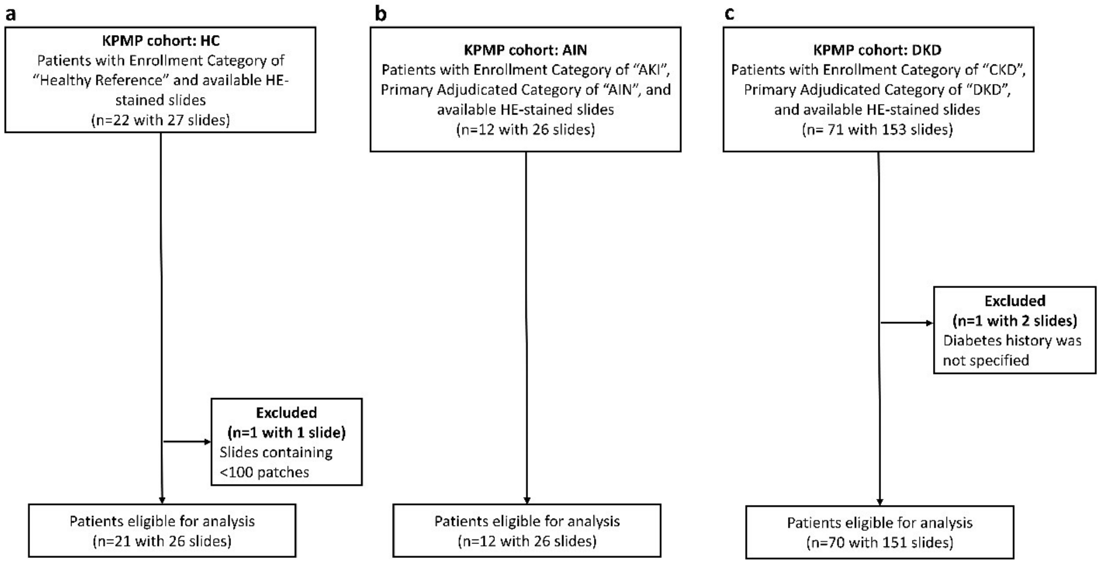


**Supplementary Figure S1. Flowcharts for patients' inclusion and exclusion in the KPMP dataset**

(a) For the HC group in the KPMP dataset, patients with Enrollment Category of "Healthy Reference" and available H&E-stained slides were included (n=22 with 27 slides) and further assessed for eligibility according to the exclusion criteria. (b) For the AIN group in the KPMP dataset, patients with Enrollment Category of "AKI", Primary Adjudicated Category of "AIN", and available H&E-stained slides were included (n=12 with 26 slides) and further assessed for eligibility according to the exclusion criteria. (c) For the DKD group in the KPMP dataset, patients with Enrollment Category of "CKD", Primary Adjudicated Category of "DKD", and available H&E-stained slides were included (n=71 with 153 slides) and further assessed for eligibility according to the exclusion criteria.

Supplementary Figure S2

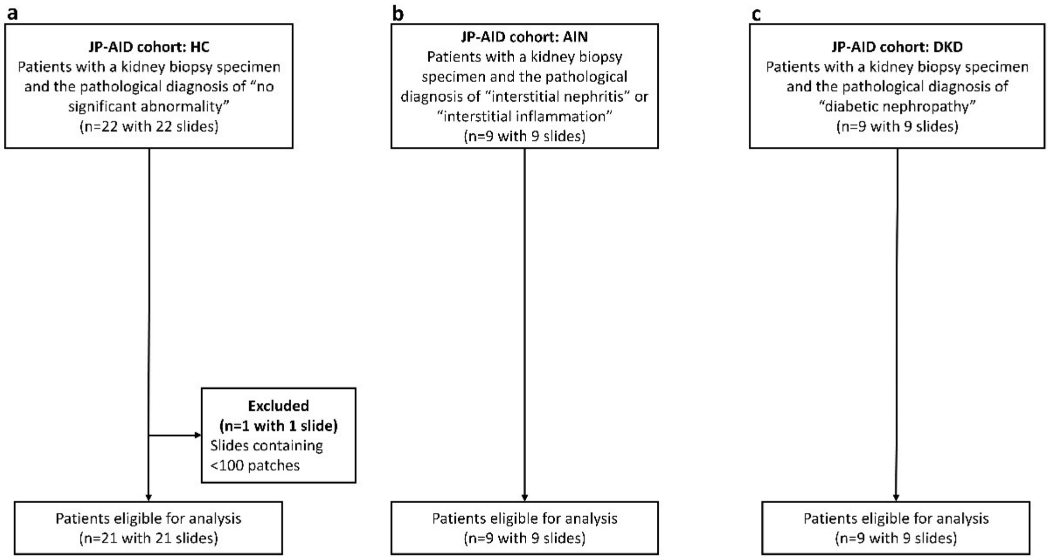


**Supplementary Figure S2. Flowcharts for patients' inclusion and exclusion in the JP-AID dataset**

(a) For the HC group in the JP-AID dataset, patients with a kidney biopsy specimen and the pathological diagnosis of "no significant abnormality" were included (n=22 with 22 slides) and further assessed for eligibility according to the exclusion criteria. (b) For the AIN group in the JP-AID dataset, patients with a kidney biopsy specimen and the pathological diagnosis of "interstitial nephritis" or "interstitial inflammation" were included (n=9 with 9 slides) and further assessed for eligibility according to the exclusion criteria. (c) For the DKD group in the JP-AID dataset, patients with a kidney biopsy specimen and the pathological diagnosis of "diabetic nephropathy" were included (n=9 with 9 slides) and further assessed for eligibility according to the exclusion criteria.

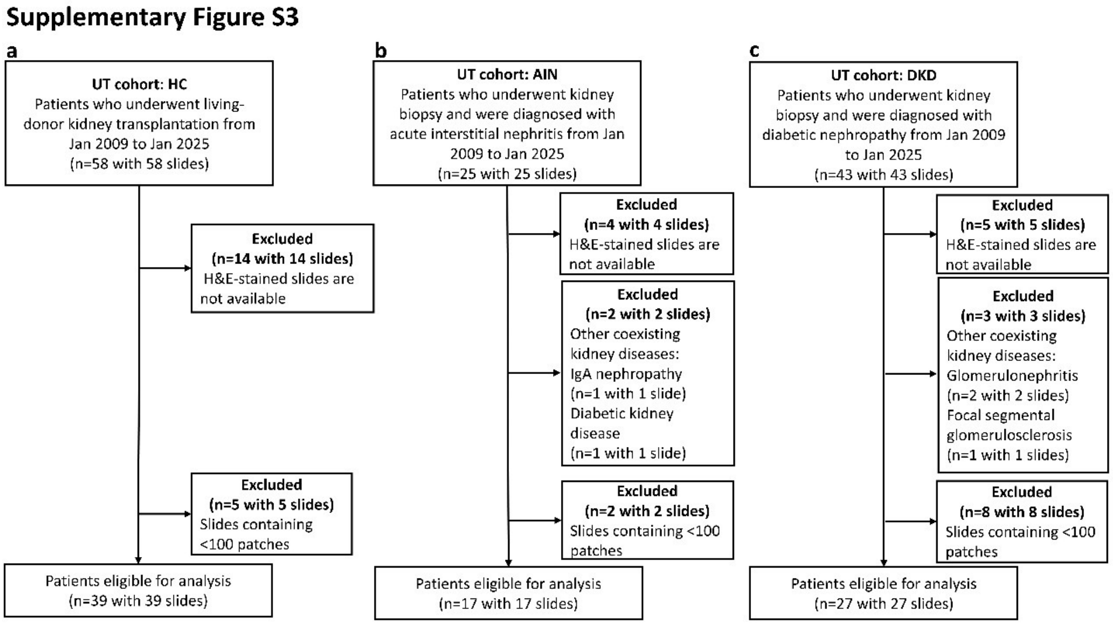


**Supplementary Figure S3. Flowcharts for patients' inclusion and exclusion in the UT dataset**

(a) For the HC group in the UT dataset, patients who underwent living-donor kidney transplantation between January 2009 and January 2025 at the University of Tokyo Hospital were included (n=58 with 58 slides) and further assessed for eligibility according to the exclusion criteria. (b) For the AIN group in the UT dataset, patients who underwent kidney biopsy and were diagnosed with AIN between January 2009 and January 2025 at the University of Tokyo Hospital were included (n=25) and further assessed for eligibility according to the exclusion criteria. (c) For the DKD group of the UT dataset, patients who underwent kidney biopsy and were diagnosed with diabetic kidney disease between January 2009 and January 2025 at the University of Tokyo Hospital were included (n=43) and further assessed for eligibility according to the exclusion criteria.

**Supplementary Figure S4**

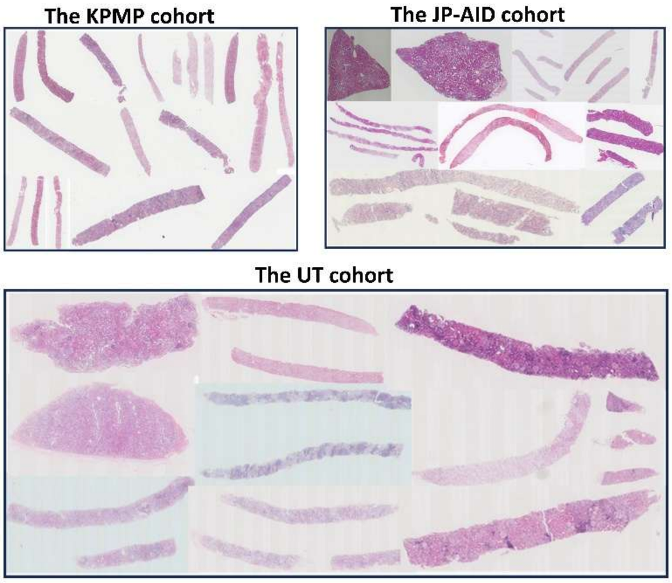


**Supplementary Figure S4. Thumbnails of example slides form each dataset**

**Supplementary Table S1. Hyperparameters for MIL models**

| model | Max pooling | ABMIL | TransMIL | CLAM |
|---|---|---|---|---|
| batch_size | 1 | 64 | 64 | 1 |
| optimizer | adam | adam | adam | adam |
| learning rate | 1e-4 | 1e-4 | 1e-4 | 1e-4 |
| weight decay | 1e-5 | 1e-5 | 1e-5 | 1e-5 |
| loss | cross entropy | cross entropy | cross entropy | cross entropy |
| save_monitor | valid_loss | valid_loss | valid_loss | valid_loss |
| epochs | 50 | 50 | 50 | 50 |
| output_shape | 3 | 3 | 3 | 3 |
| bag_weight | | | | 0.7 |
| dropout | | | | False |
| k_sample | | | | 8 |

ABMIL, attention-based multiple instance learning; TransMIL, transformer-based multiple instance learning: CLAM, clustering-constraint attention multiple instance learning.

#### Supplementary methods

##### Dataset description

For the development cohort, we employed publicly available datasets from the Kidney Precision Medicine Project (KPMP, accessed on 20th January 2025) and the Japan-Pathology Artificial Intelligence Diagnostics Project (JP-AID, accessed on 20th January 2025). KPMP is a multi-year project funded by National Institute of Diabetes and Digestive and Kidney Diseases (NIDDK) to better understand the mechanisms of chronic kidney disease (CKD) and acute kidney injury (AKI) using a tissue centric, research biopsy driven approach. KPMP establishes a publicly available dataset, including WSIs. Formalin-fixed paraffin-embedded (FFPE) H&E-stained WSIs from the KPMP repository were used to develop the model. The KPMP dataset comprised 26 slides from 21 HCs, 26 slides from 12 patients with AIN, and 151 slides from 70 patients with DKD. HC cases were selected based on the Enrollment Category of "Healthy Reference". Among the 21 HC cases from KPMP, 2 were obtained via nephrectomy, and remaining were derived from pre-perfusion biopsies from transplanted kidneys. AIN cases were selected from those with the Enrollment Category of "AKI" and the Primary Adjudicated Category of "AIN". DKD cases were selected from those with the Enrollment Category of "CKD" and the Primary Adjudicated Category of "DKD". The JP-AID is a multi-institutional project aimed at developing and implementing AI tools for pathology diagnostic support conducted by Japanese Society of Pathology. JP-AID database comprises approximately 10000 WSIs gathered from 13 institutions in Japan. From the JP-AID database, we collected H&E-stained WSIs comprising 21 HC slides, of which 20 were derived from post-transplant cases, nine slides of DKD, and nine slides of AIN. Cases from the JP-AID database were selected based on the following criteria. HC cases were included if the organ was "kidney" and the pathological diagnosis was "no significant abnormality". AIN cases were selected if the organ was "kidney" and the pathological diagnosis was "interstitial nephritis" or "interstitial inflammation". DKD cases were identified if the organ was "kidney" and the pathological diagnosis was "diabetic nephropathy". Data from the JP-AID database were anonymized, and detailed clinical information other than age, sex, and diagnosis was not available. Of note, the demographic information for the transplanted kidneys in the JP-AID HC group may originate from recipients rather than donors. As the database does not provide detailed clarification, this remains uncertain. For external validation, an independent dataset of H&E-stained WSIs was obtained from the University of Tokyo Hospital (the UT dataset). The UT dataset included 39 slides of HC, 17 slides of AIN, and 30 slides of DKD. For HC, we collected slides of kidney specimens from living kidney donors obtained zero or one hour after transplantation from January 2009 to January 2025. For AIN or DKD, we included patients who underwent kidney biopsy in the Department of Nephrology and Endocrinology of the University of Tokyo Hospital and were diagnosed with AIN or DKD between January 2009 and January 2025. The final diagnosis was based on kidney biopsy reports. Patients with other coexisting kidney diseases (e.g. glomerulonephritis, focal segmental glomerulosclerosis, and IgA nephropathy) were excluded. For the JP-AID and UT datasets, one slide per one patient was used. This study protocol adhered to the Declaration of Helsinki and was approved by the University of Tokyo Institutional Review Board (approval number: 2024526NI).

All the WSIs from KPMP were digitized using Aperio AT2 (Leica Biosystems) at 40x magnification and provided in SVS format, and those from the University of Tokyo hospital biopsies were scanned using NanoZoomer (Hamamatsu Photonics) with 40x magnification in NDPI format. Although slides from the JP-AID dataset were available in NDPI or BIF format at magnifications of 20x or 40x, detailed information regarding the scanning method was not provided.

### TRIPOD Checklist: Prediction Model Development and Validation

| Section/Topic | Item | | Checklist Item | Page |
|---|---|---|---|---|
| **Title and abstract** | | | | |
| Title | 1 | D;V | Identify the study as developing and/or validating a multivariable prediction model, the target population, and the outcome to be predicted. | 1 |
| Abstract | 2 | D;V | Provide a summary of objectives, study design, setting, participants, sample size, predictors, outcome, statistical analysis, results, and conclusions. | 2 |
| **Introduction** | | | | |
| Background and objectives | 3a | D;V | Explain the medical context (including whether diagnostic or prognostic) and rationale for developing or validating the multivariable prediction model, including references to existing models. | 4-6 |
| | 3b | D;V | Specify the objectives, including whether the study describes the development or validation of the model or both. | 4-6 |
| **Methods** | | | | |
| Source of data | 4a | D;V | Describe the study design or source of data (e.g., randomized trial, cohort, or registry data), separately for the development and validation data sets, if applicable. | 6,7 |
| | 4b | D;V | Specify the key study dates, including start of accrual; end of accrual; and, if applicable, end of follow-up. | 6,7 |
| Participants | 5a | D;V | Specify key elements of the study setting (e.g., primary care, secondary care, general population) including number and location of centres. | 6,7 |
| | 5b | D;V | Describe eligibility criteria for participants. | 6,7 |
| | 5c | D;V | Give details of treatments received, if relevant. | NA |
| Outcome | 6a | D;V | Clearly define the outcome that is predicted by the prediction model, including how and when assessed. | 6,7 |
| | 6b | D;V | Report any actions to blind assessment of the outcome to be predicted. | NA |
| Predictors | 7a | D;V | Clearly define all predictors used in developing or validating the multivariable prediction model, including how and when they were measured. | NA |
| | 7b | D;V | Report any actions to blind assessment of predictors for the outcome and other predictors. | NA |
| Sample size | 8 | D;V | Explain how the study size was arrived at. | NA |
| Missing data | 9 | D;V | Describe how missing data were handled (e.g., complete-case analysis, single imputation, multiple imputation) with details of any imputation method. | NA |
| Statistical analysis methods | 10a | D | Describe how predictors were handled in the analyses. | NA |
| | 10b | D | Specify type of model, all model-building procedures (including any predictor selection), and method for internal validation. | 8-11 |
| | 10c | V | For validation, describe how the predictions were calculated. | 8-11 |
| | 10d | D;V | Specify all measures used to assess model performance and, if relevant, to compare multiple models. | 11 |
| | 10e | V | Describe any model updating (e.g., recalibration) arising from the validation, if done. | NA |
| Risk groups | 11 | D;V | Provide details on how risk groups were created, if done. | NA |
| Development vs. validation | 12 | V | For validation, identify any differences from the development data in setting, eligibility criteria, outcome, and predictors. | 6,7 |
| **Results** | | | | |
| Participants | 13a | D;V | Describe the flow of participants through the study, including the number of participants with and without the outcome and, if applicable, a summary of the follow-up time. A diagram may be helpful. | 12-14 |
| | 13b | D;V | Describe the characteristics of the participants (basic demographics, clinical features, available predictors), including the number of participants with missing data for predictors and outcome. | 12-14 |
| | 13c | V | For validation, show a comparison with the development data of the distribution of important variables (demographics, predictors and outcome). | 13,14 |
| Model development | 14a | D | Specify the number of participants and outcome events in each analysis. | 12,13 |
| | 14b | D | If done, report the unadjusted association between each candidate predictor and outcome. | NA |
| Model specification | 15a | D | Present the full prediction model to allow predictions for individuals (i.e., all regression coefficients, and model intercept or baseline survival at a given time point). | NA |
| | 15b | D | Explain how to the use the prediction model. | 13,14 |
| Model performance | 16 | D;V | Report performance measures (with CIs) for the prediction model. | 13,14 |
| Model-updating | 17 | V | If done, report the results from any model updating (i.e., model specification, model performance). | NA |
| **Discussion** | | | | |
| Limitations | 18 | D;V | Discuss any limitations of the study (such as nonrepresentative sample, few events per predictor, missing data). | 19 |
| Interpretation | 19a | V | For validation, discuss the results with reference to performance in the development data, and any other validation data. | NA |
| | 19b | D;V | Give an overall interpretation of the results, considering objectives, limitations, results from similar studies, and other relevant evidence. | 16-20 |
| Implications | 20 | D;V | Discuss the potential clinical use of the model and implications for future research. | 19,20 |
| **Other information** | | | | |
| Supplementary information | 21 | D;V | Provide information about the availability of supplementary resources, such as study protocol, Web calculator, and data sets. | 21-23 |
| Funding | 22 | D;V | Give the source of funding and the role of the funders for the present study. | 22 |

*Items relevant only to the development of a prediction model are denoted by D, items relating solely to a validation of a prediction model are denoted by V, and items relating to both are denoted D;V. We recommend using the TRIPOD Checklist in conjunction with the TRIPOD Explanation and Elaboration document.
